# Antibody responses against SARS-CoV-2 variants induced by four different SARS-CoV-2 vaccines

**DOI:** 10.1101/2021.09.27.21264163

**Authors:** Marit J. van Gils, A. H. Ayesha Lavell, Karlijn van der Straten, Brent Appelman, Ilja Bontjer, Meliawati Poniman, Judith A. Burger, Melissa Oomen, Joey H. Bouhuijs, Lonneke A. van Vught, Marleen A. Slim, Michiel Schinkel, Elke Wynberg, Hugo D.G. van Willigen, Marloes Grobben, Khadija Tejjani, Jacqueline van Rijswijk, Jonne L. Snitselaar, Tom G. Caniels, Amsterdam UMC COVID-19 S3/HCW study group, Alexander P. J. Vlaar, Maria Prins, Menno D. de Jong, Godelieve J. de Bree, Jonne J. Sikkens, Marije K. Bomers, Rogier W. Sanders

## Abstract

**Background:** Emerging and future SARS-CoV-2 variants may jeopardize the effectiveness of vaccination campaigns. Therefore, it is important to know how the different vaccines perform against diverse SARS-CoV-2 variants.

**Methods:** In a prospective cohort of 165 SARS-CoV-2 naive health care workers, vaccinated with either one of four vaccines (BNT162b2, mRNA-1273, AZD1222 or Ad26.COV2.S), we performed a head-to-head comparison of the ability of sera to recognize and neutralize SARS-CoV-2 variants of concern (VOCs; Alpha, Beta, Gamma, Delta and Omicron). Repeated serum sampling was performed 5 times during a year (from January 2021 till January 2022), including before and after booster vaccination with BNT162b2.

**Findings:** Four weeks after completing the initial vaccination series, SARS-CoV-2 wild-type neutralizing antibody titers were highest in recipients of BNT162b2 and mRNA-1273 (geometric mean titers (GMT) of 197 [95% CI 149-260] and 313 [95% CI 218-448], respectively), and substantially lower in those vaccinated with the adenovirus vector-based vaccines AZD1222 and Ad26.COV2.S (GMT of 26 [95% CI 18-37] and 14 [95% CI 8-25] IU/ml, respectively). These findings were robust for adjustment to age and sex. VOCs neutralization was reduced in all vaccine groups, with the largest (9- to 80-fold) reduction in neutralization observed against the Omicron variant. The booster BNT162b2 vaccination increased neutralizing antibody titers for all groups with substantial improvement against the VOCs including the Omicron variant. Study limitations include the lack of cellular immunity data.

**Conclusions:** Overall, this study shows that the mRNA vaccines appear superior to adenovirus vector-based vaccines in inducing neutralizing antibodies against VOCs four weeks after initial vaccination and after booster vaccination.

## Introduction

As of March 2022, the coronavirus disease 2019 (COVID-19) pandemic has caused over 458 million confirmed infections and over 6 million reported deaths [1], calling for strong interventions. A number of vaccines have been developed that proved efficacious in preventing severe acute respiratory syndrome coronavirus 2 (SARS-CoV-2) infection, the causative agent of COVID-19, and/or severe disease from infection, providing hope that we can halt this pandemic. Three vaccines, i.e. those developed by Pfizer-BioNTech (BNT162b2/Comirnaty), Moderna (mRNA-1273/Spikevax) and J&J/Janssen (Ad26.COV2.S), have been approved (for emergency use) in the United States by the FDA, while the EMA in the European Union has additionally approved (for emergency use) a fourth vaccine from Oxford/AstraZeneca (AZD1222/Vaxzevria), and very recently a fifth from Novavax (NVX-CoV2372/Nuvaxovid). Early efficacy trials showed that the mRNA vaccines BNT162b2 and mRNA-1273 had high efficacy (>90%) against symptomatic infection, whereas the adenovirus vector-based vaccines AZD1222 and Ad26.COV2.S resulted in lower vaccine efficacy (60-70%) against symptomatic infection [2–5]. Efficacy waned somewhat over time for all vaccines [6]. However, all vaccines were extremely effective at preventing severe disease. Neutralizing antibodies proved to be a very strong correlate of protection [7–9]. So far, over 10.7 billion COVID-19 vaccine doses have been administered worldwide [1].

Since the start of the pandemic, SARS-CoV-2 has diversified considerably, both genetically and antigenically. Currently, five virus lineages have been designated as a variant of concern (VOC) by the WHO due to, among others, suspected increased transmissibility or virulence: Alpha (B.1.1.7/20I/N501Y.V1), Beta (B.1.351/20H/N501Y.V2), Gamma (B.1.1.28.P1/P.1/20J/N501Y.V3), Delta (B.1.617.2/21A) and Omicron (B.1.1.529/21K/BA.1). All five VOCs have spread globally, but only Delta and Omicron are currently circulating with Omicron being the dominant variant [11]. In addition to the five VOCs, the WHO has defined a number of variants of interest (VOIs) that should be monitored closely as well. From studies on monoclonal antibodies, including ones developed for therapeutic application in COVID-19, it is known that these can lose neutralization potency against the VOCs and VOIs, in particular those targeting the receptor binding motive (RBM) on the SARS-CoV-2 spike (S) protein [12]. The most relevant mutations for loss of neutralization in Alpha, Beta, Gamma and Delta include E484K, K417T/N and L452R/Q in the receptor binding domain (RBD) and Δ69-70 and Δ242-244 in the N-terminal domain (NTD), while Omicron has many more mutations: 32 in S, including 15 in RBD. Considering the pandemic is still ongoing, it is important to know how the different vaccines perform against the different SARS-CoV-2 variants.

In randomized clinical trials and real-world observational studies, several vaccines proved less efficacious against VOCs, in particular the Beta and Delta variants [13–20]. In England, reduced effectiveness against symptomatic COVID-19 was observed with the Delta variant compared to the Alpha variant [21], in particular after a single vaccine dose. The emerging data indicates that vaccine efficacy is further and substantially reduced against Omicron, necessitating booster immunizations [22–24]. In line with these observations, VOCs were shown to be less sensitive to neutralizing antibodies induced by infection or vaccination. Antibody responses are generally sufficient to neutralize the Alpha variant to similar levels as the original Wuhan strain in mRNA vaccine recipients and in convalescent individuals. However, the Beta, Gamma, Delta and Omicron variants showed on average a 9-fold, 4-fold, 4-fold and 20-to 40-fold reduced sensitivity respectively to neutralization by sera from convalescent patients as well as from vaccine recipients [25–27].

Although previous studies have provided valuable initial insights in the sensitivity of VOCs to neutralization induced by infection or vaccination, few studies have directly compared the ability of humoral responses induced by the four different vaccines to cope with VOCs. Previous studies have used diverse serological assays, mainly focused on one or two vaccines, or used regression models to combine studies, complicating direct comparisons. Here, we present a head-to-head comparison of the binding and neutralizing activity against the VOCs in serum of individuals after the initial vaccination series with BNT162b2, mRNA-1273, AZD1222 or Ad.COV2.S vaccine and subsequently after a BNT162b booster vaccination 5-11 months later.

## Methods and Materials

### Study design

Since March 2020, we followed a cohort of hospital health care workers (HCW) in the Amsterdam University Medical Centers, consisting of two tertiary care hospitals (S3 study, Netherlands Trial Register NL8645) [28]. This study is reported as per the Strengthening the Reporting of Observational Studies in Epidemiology (STROBE) guideline. Participants underwent frequent phlebotomies to determine seroconversion against SARS-CoV-2, measured by total Ig against S1-RBD using enzyme-linked immunosorbent assay (Wantai ELISA). Between January and May 2021 participants of the cohort were vaccinated with either BNT162b2, mRNA-1273, AZD1222, or a single dose Ad.26CoV2.S (depending on the national distribution of available vaccines). Blood samples were taken approximately three weeks after the first vaccine with BNT162b2, mRNA-1273 and AZD1222 and four weeks after the second vaccine. In the case of vaccination with Ad.26CoV2.S, blood samples were taken approximately four to five and eight weeks after vaccination (Figure 1A). Preferably a blood sample was taken within days before the first vaccine was administered. Only seronegative HCW were included in the analysis. Between October 2021 and January 2022, the cohort was again invited for serum collection before and after BNT162b2 booster vaccination. Due to low attendance of the group vaccinated with mRNA-1273 or Ad26.COV2.S we included 16 additional SARS-CoV-2 naive HCW.

**Figure 1:**
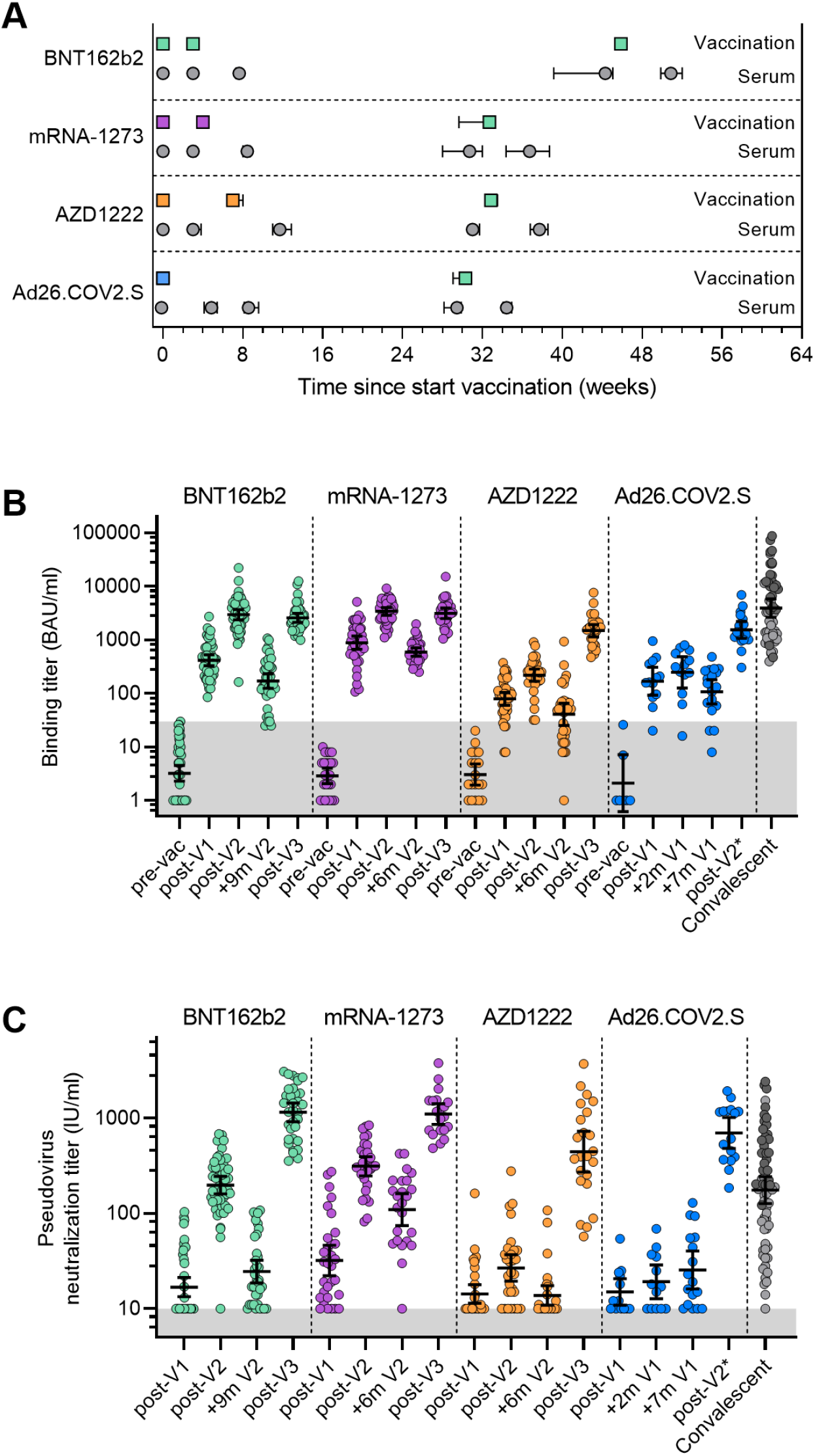
Binding and neutralization titers pre- and post-vaccination with one of the four SARS-CoV-2 vaccines. (A) Timelines of the vaccinations and serum collections, showing the mean and interquartile range of times of vaccination and samples in weeks after the first dose. (B) Binding titers to wild-type S protein (BAU/ml) of 1:100,000 diluted sera collected over time for the four vaccination groups. The convalescent group (n=67) consists of sera from hospitalized (dark gray) and non-hospitalized (light gray) COVID-19 patients collected 4-6 weeks post symptom onset. Geometric mean titers (GMT) and 95% confidence intervals (CI) are indicated. The lower cutoff for binding was set at 30 BAU/ml (grey shading). (C) Neutralization IC_50_ titers (IU/ml) of D614G pseudovirus for sera collected post-vaccination over time for the four vaccination groups. The convalescent group (n=67) consists of sera from hospitalized (dark gray) and non-hospitalized (light gray) COVID-19 patients collected 4-6 weeks post symptom onset. GMT and 95% CI are indicated. The lower cutoff for neutralization was set at an IC_50_ of 10 or for Omicron at 2 IU/ml (grey shading). * As the Ad26.COV2.S vaccine uses a single-dose regime, the BNT162b2 booster vaccination is vaccination number two (V2). All data points shown here represent the mean of a technical triplicate. Uni- and multivariable linear regression analysis results in Table S4.

To enable comparison between antibody response following vaccination and following infection, we included serum from two COVID-19 patient cohorts. Participants in the COSCA cohort were included from March 2020 till the end of January 2021, with the wild-type and D614G variant being the dominant circulating strains [29]. These include hospitalized and non-hospitalized participants and serum was obtained four to six weeks after symptom onset [30]. A serum pool was created from COSCA samples of 68 participants. Another serum pool was created from sera collected in the RECoVERED cohort [31]. In total, 251 RECoVERED serum samples were used, obtained up to seven months post start of symptoms (median of 3 months) from participants who experienced mild, moderate or severe COVID-19. The S3 study, the COSCA study and the RECoVERED study were approved by the medical ethical review board of the Amsterdam University Medical Centers (NL73478.029.20, NL73281.018.20 and NL73759.018.20, respectively). All participants provided written informed consent.

### Protein design

The mutations compared to the WT variant (Wuhan Hu-1; GenBank: MN908947.3) in the S proteins are depicted in Table S1. The S constructs were ordered as gBlock gene fragments (Integrated DNA Technologies) and cloned in a pPPI4 expression vector containing a hexahistidine (his) tag with Gibson Assembly (ThermoFisher) [32]. All S constructs were verified by Sanger sequencing, subsequently produced in HEK293F cells (ThermoFisher), and purified as previously described [32].

### Protein coupling to Luminex beads

To measure the binding of IgG to the spike proteins of different VOCs, we covalently coupled pre-fusion stabilized spike proteins to Luminex Magplex beads using a two-step carbodiimide reaction as previously described [33]. In short, Luminex Magplex beads (Luminex) were washed with 100 mM monobasic sodium phosphate pH 6.2 and activated by addition of Sulfo-N-Hydroxysulfosuccinimide (Thermo Fisher Scientific) and 1-Ethyl-3-(3-dimethylaminopropyl) carbodiimide (Thermo Fisher Scientific) and incubated for 30 minutes on a rotator at room temperature. After washing the activated beads three times with 50 mM MES pH 5.0, the spike proteins were added in ratio of 75 µg protein to 12.5 million beads and incubated for three hours on a rotator at room temperature. To block the beads for aspecific binding, we incubated the beads for 30 minutes with PBS containing 2% BSA, 3% fetal calf serum and 0.02% Tween-20 at pH 7.0. Finally, the beads were washed and stored at 4°C in PBS containing 0.05% sodium azide.

### Luminex assays

Optimization experiments determined the optimal concentration of the sera for studying the humoral vaccination response to be 100.000-fold dilution. As previously described [34], 50 µL of a bead mixture containing all different spike proteins in a concentration of 20 beads per µL were added to 50 µL of diluted serum and incubated overnight on a rotator at 4°C. The next day, plates were washed with TBS containing 0.05% Tween-20 (TBST) and resuspended in 50 µL of Goat-anti-human IgG-PE (Southern Biotech). After 2 hours of incubation on a rotator at room temperature, the beads were washed with TBST and resuspended in 70 µL Magpix drive fluid (Luminex). Read-out of the plates was performed on a Magpix (Luminex). The binding of antibodies was determined as the Median Fluorescence Intensity (MFI) of approximately 50 to 100 beads per well, corrected for background signals by subtracting the MFI of wells containing only buffer and beads and converted into binding antibody units per ml (BAU/ml) using the WHO International Standard for anti-SARS-CoV-2 immunoglobulin (NIBSC 20/136). To confirm assay performance, a titration of serum of one convalescent COVID-19 patient as well as positive and negative controls were included on each plate. In addition, 15 to 20% of samples of each run were replicated to confirm the results.

### Pseudovirus construction

The WT, D614G, Alpha, Alpha E484K, Beta, Gamma and Omicron BA.1 pseudovirus SARS-CoV-2-S constructs were ordered as gBlock gene fragments (Integrated DNA Technologies) and cloned using SacI and ApaI in the pCR3 SARS-CoV-2-S_Δ19_ expression plasmid [35] using Gibson Assembly (ThermoFisher). Pseudovirus SARS-CoV-2-S expression constructs for Delta and Kappa were provided by Dr. Beatrice Hahn, while those for Beta Δ242-244, Lambda, Epsilon, Iota and Zeta were provided by Drs. Paul Bieniasz and Theodora Hatziioannou. All constructs were verified by Sanger sequencing and the mutations for the VOCs and VOIs are indicated in Table S1. Pseudoviruses were produced by co-transfecting the SARS-CoV-2-S expression plasmid with the pHIV-1_NL43_ ΔEnv-NanoLuc reporter virus plasmid in HEK293T cells (ATCC, CRL-11268), as previously described [35]. Cell supernatant containing the pseudovirus was harvested 48 hours post transfection and stored at −80°C until further use.

### Pseudovirus neutralization assay

Neutralization activity was tested using a pseudovirus neutralization assay, as previously described[32]. Shortly, HEK293T/ACE2 cells, kindly provided by Dr. Paul Bieniasz [35], were seeded at a density of 20,000 cells/well in a 96-well plate coated with 50 μg/mL poly-L-lysine one day prior to the start of the neutralization assay. Heat-inactivated sera samples were serially diluted in cell culture medium (DMEM (Gibco), supplemented with 10% FBS, penicillin (100 U/mL), streptomycin (100 μg/mL) and GlutaMax (Gibco)), mixed in a 1:1 ratio with pseudovirus and incubated for 1 hour at 37°C. Subsequently, these mixtures were added to the cells in a 1:1 ratio and incubated for 48 hours at 37°C, followed by a PBS wash and lysis buffer to measure the luciferase activity in cell lysates using the Nano-Glo Luciferase Assay System (Promega) and GloMax system (Turner BioSystems). Relative luminescence units (RLU) were normalized to the positive control wells where cells were infected with pseudovirus in the absence of NAbs or sera. The neutralization titers (IC_50_) were determined as the serum dilution or antibody concentration at which infectivity was inhibited by 50%, respectively, using a non-linear regression curve fit (GraphPad Prism software version 8.3) and serum dilutions were converted into international units per ml (IU/ml) using the WHO International Standard for anti-SARS-CoV-2 immunoglobulin (NIBSC 20/136). Samples with virus neutralization titers of <10 IU/ml were defined as having undetectable neutralization. Neutralization titers from this pseudovirus assay have been shown to strongly correlate with titers obtained in an authentic virus neutralization assay [32].

### Statistical analysis

We used univariable and multivariable linear regression analysis to compare antibody binding or neutralization titers between subjects. We used univariable and multivariable linear mixed model analysis with a random intercept to compare changes in antibody binding or neutralization titers in analyses that included comparisons within subjects. Two-sided p-values <0.05 were considered significant. Outcomes were log-transformed before analysis. All multivariable models comprised age and sex, whereas the comparisons pre-booster vaccination also included the time since previous vaccination as variable. All regression analyses were performed in R (version 4.0.3), using the lme4 package for mixed models. Spearman’s rank correlation was performed for the comparison between median neutralization titer per vaccine group and reported vaccine efficacy. Data visualization and Spearman’s rank correlation analyses were performed in GraphPad Prism software (version 8.3). The reported vaccine efficacy data has been taken from current literature (Table S2) [2,3,5,14–18,20,22–24,36–52]. After initial submission, the initial preplanned analysis plan was extended with multivariable and mixed model analyses because of the addition of booster vaccine data and suggestions of reviewers.

## Results

### Binding and neutralizing antibody responses after initial vaccination series

In a direct head-to-head comparison, using the same assays, we assessed the ability of four FDA and/or EMA approved SARS-CoV-2 vaccines to induce humoral immune responses in humans. From the S3 HCW cohort [53], we included SARS-CoV-2 naive individuals who completed BNT162b2 (n=54), mRNA-1273 (n=43), AZD1222 (n=42) or Ad26.COV2.S vaccination (n=26; Table S3) and received a BNT162b2 booster vaccination. Although the four vaccine groups were fairly similar in composition, 64.8-86.0% female with the majority between 35-60 years old (Table 1), the AZD1222 group mostly consists of individuals over 60 years of age, because the Dutch government restricted the use of AZD1222 to this age group due to safety concerns. Furthermore, the Ad26.COV2.S group included fewer individuals because the Dutch government temporarily restricted its use because of similar reasons [54]. For vaccinees who received the BNT162b2, mRNA-1273 and AZD122 vaccines, samples were taken approximately three weeks after the first vaccination and four weeks after the second vaccination (Figure 1A). As the Ad26.COV2.S vaccine uses a single-dose regime, vaccine recipients were sampled approximately five and eight weeks after the single-dose vaccination. In addition, serum samples were collected pre- and four weeks post-BNT162b2 booster vaccination.

**Table 1.**
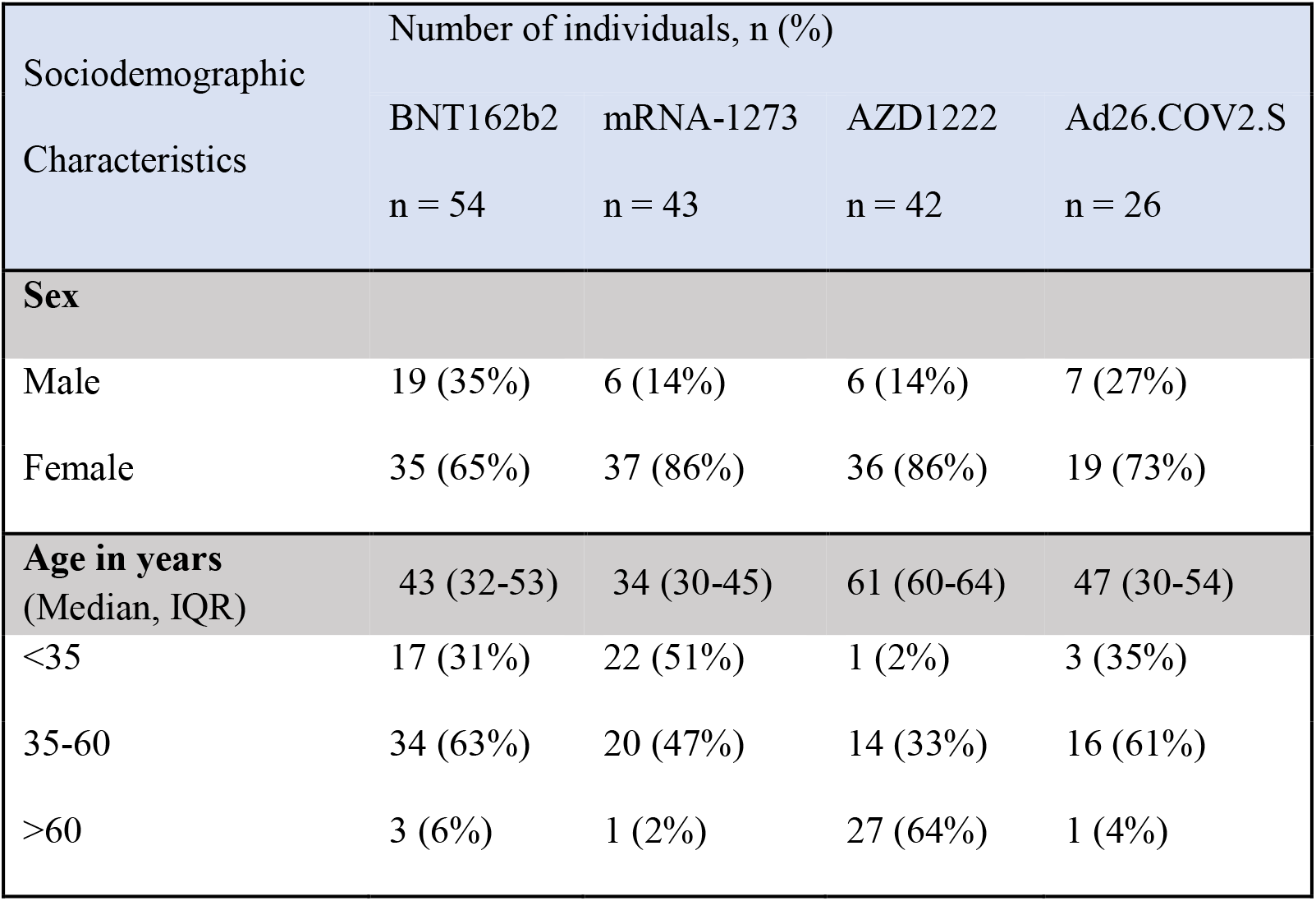
Sociodemographic characteristics.

We first assessed S protein binding titers in vaccinee sera after the initial vaccination series in a custom luminex assay against the wild-type (WT) S protein. Overall, the antibody responses against the S protein were relatively homogeneous within each group, showing larger intergroup than intragroup difference, with only one Ad26.COV2.S recipient having binding titers below the limit of detection after the initial vaccination series (Figure 1B, Table S5). Antibody responses in fully vaccinated mRNA-1273 and BNT162b2 recipients were comparable with convalescent individuals 4-6 weeks after symptom onset (COSCA study, n = 68) (geometric mean titer [GMT] of 3461 [95% CI 2472-4847] and 2967 [95% CI 2212-3979] versus 3463 [95% CI 2630-4559] BAU/ml, respectively), but the responses of AZD1222 and Ad26.COV2.S recipients were substantially (16 to 20-fold) lower (GMT of 219 [95% CI 153-313] and 169 [95% CI 95-300] BAU/ml, respectively). The difference in GMT between mRNA and vector vaccines did not significantly change after correcting for age and sex as possible confounders (Table S4).

Next, we tested the neutralizing activity of vaccinee sera using a lentiviral-based pseudovirus assay of the SARS-CoV-2 D614G (B.1) variant (Figure 1C, Table S5). We detected the highest neutralization activity in mRNA-1273 recipients, followed by BNT162b2 recipients and convalescent individuals (GMT IC_50_ of 313 [95% CI 218-448] versus 197 [95% CI 149-260] and 175 [95% CI 138-223] IU/ml, respectively), and approximately an order of magnitude lower activity in those vaccinated with the adenovirus vector-based vaccines AZD1222 and Ad26.COV2.S (GMT of 26 [95% CI 18-37] and 14 [95% CI 8-25] IU/ml, respectively). In 1/50 BNT162b2, 7/30 AZD1222 and 6/13 Ad26.COV2.S recipients there was no detectable neutralization activity (IC_50_ < 10 IU/ml) (Figure 1C, Table S4, S5). The neutralization titers remained significantly different between the two mRNA vaccine groups, and between the mRNA and vector vaccinees, after correcting for age and sex as possible confounders (Table S4). The differences in humoral immune responses between the groups following vaccination are consistent with the reported observed differences in the efficacy of these vaccines (Table S2) and in agreement with the observations that neutralizing antibodies are a strong correlate of protection [19,55,56].

### Binding and neutralizing antibody responses after a single vaccination

We also assessed the responses after a single vaccination of one of four vaccines. First, we wished to directly compare the single-dose of Ad26.COV2.S with one dose of each of the other three vaccines. Second, we wanted to gauge the level of humoral immunity after partial vaccination, which is relevant when vaccinating during an infection wave and/or when considering to postpone the second vaccination.

All BNT162b2 and mRNA-1273 recipients had detectable antibody binding titers against S after one vaccination, while 5 out of 42 AZD1222 and 1 of 13 Ad26.COV2.S recipients did not (Figure 1B, Table S5). The binding antibody titers were highest for the mRNA vaccine groups with mRNA-1273 recipients (GMT 781 [95% CI 512-1191] BAU/ml) exceeding not only the level after one vaccination of the three other vaccines (GMT 394 [95% CI 280-554], 95 [95% CI 60-150] and 169 [95% CI 96-296] BAU/ml for BNT162b2, AZD1222 and Ad26.COV2.S, respectively; corrected for age and sex), but also the binding titers after two doses of AZD1222 (GMT 202 [95% CI 136-301] BAU/ml). The neutralizing antibody levels after one dose were low in all cases (GMT 15 [95% CI 10-20] IU/ml for BNT162b2, GMT 13 [95% CI 8-22] IU/ml for AZD1222, GMT 13 [95% CI 8-23] IU/ml for Ad26.COV2.S and GMT 28 [95% CI 18-42] IU/ml for mRNA-1273; corrected for age and sex) with only 19 of 45 (42%) BNT162b2, 13 of 35 (37%) AZD1222, 7 of 13 (54%) Ad26.COV2.S and 26 of 31 (84%) mRNA-1273 having detectable neutralization (IC_50_ >10 IU/ml) (Figure 1C, Table S4, S5). Furthermore, eight weeks after the single Ad26.COV2.S vaccination the neutralization titers were slightly increased compared to the five week samples (GMT IC_50_ of 13 versus 17 IU/ml), although not significant, and two additional recipients showed detectable neutralization indicative of some maturation of the antibody response.

### Binding and neutralizing antibody responses pre- and post-booster vaccination

To study the decline of antibody binding and neutralization levels, serum samples were collected 5-11 months post vaccination, just prior to the BNT162b2 booster vaccination. We observed significant antibody binding titer decline rates for all vaccine groups except the Ad26.COV2.S group, which showed no decline between four weeks post initial vaccination series and pre-booster vaccination (fold change 0.47 [95% 0.19-1.18] corrected for age, sex and time; Figure 1B, S1A and Table S4, S5). Similarly, decline rates of neutralizing antibody titers were also significant for BNT162b2, mRNA-1273 and AZD1222 recipients (fold change 0.13 [95% CI 0.10-0.18], 0.36 [95% CI 0.25-0.52] and 0.33 [95% CI 0.23-0.47], respectively), but not for Ad26.COV2.S recipients (fold change 1.08 [95% CI 0.59-1.95]; corrected for age, sex and time; Figure 1C, S1B, Table S4, S5).

After the BNT162b2 booster vaccination, all groups showed a significant increase in antibody binding and neutralization titers with all participants showing detectable neutralization. The binding antibody levels post-booster were significantly lower for individuals originally vaccinated with AZD1222 or Ad26.COV2.S (GMT 1480 [95% CI 997-2196] and 1484 [95% CI 1020-2140], respectively) compared to those vaccinated with mRNA-1273 or BNT162b2 (GMT 2993 [95% CI 2097-4271] and 2756 [95% CI 2071-3668], respectively), while this difference was not seen for neutralizing antibody levels (Figure 1A-B, Table S4). The fold increase of neutralization titers pre- and post-booster was less for individuals originally vaccinated with mRNA-1273 (fold change 9.74 [95% CI 6.62-14.34] corrected for age and sex), compared to BNT162b2, AZD1222 or Ad26.COV2.S recipients (fold change 46.85 [95% CI 34.50-63.68], 54.54 [95% 38.11-77.96] and 33.79 [95% CI 21.72-52.56], respectively; corrected for age and sex). Still, the neutralization titers were the lowest for those originally vaccinated with AZD1222 (GMT 379 [95% CI 221-650] IU/ml, corrected for age and sex) compared to those vaccinated with BNT162b2, mRNA-1273 or Ad26.COV2.S (GMT 1192 [95% CI 817-1738], 1160 [95% CI 717-1878] and 735 [95% CI 445-1213] IU/ml, respectively; Figure 1A-B, Table S4, S5). When compared to titers after the initial vaccination series, the post-booster binding titers were not significantly different for BNT162b2 and mRNA-1273 recipients (fold change 0.98 [95% CI 0.61-1.58] and 0.94 [95% CI 0.54-1.62], respectively; corrected for age and sex), while they were significantly higher in AZD-1222 and Ad26.COV2.S recipients (fold change 6.86 [95% CI 3.85-12.20] and 6.26 [95% CI 2.76-14.15], respectively, Table S4). For all groups, neutralizing antibody levels were significantly higher post-booster compared to post initial vaccination.

### Binding and neutralizing antibody responses against VOCs

After initial vaccination series, the binding antibody responses against VOCs S proteins were similar to those against WT S protein (Figure S2A, Table S5) as was the ranking of the different vaccines. Thus, mRNA vaccine recipients had higher binding responses compared to adenovirus vector-based vaccine recipients for all VOCs.

We next tested the neutralizing activity of the vaccine sera against the five VOCs after initial vaccination series (Figure 2A, Figure S2B, Table S5). As emerging data indicates that neutralization against Omicron is substantially reduced, we tested the sera against this VOC, specifically the BA.1 variant, at lower dilutions. The neutralizing titers against the VOCs were highest in the mRNA recipients (GMT IC_50_ of 116 (Alpha), 49 (Beta), 103 (Gamma), 65 (Delta), 2 IU/ml Omicron for BNT162b2 recipients, and 201 (Alpha), 68 (Beta), 225 (Gamma), 155 (Delta), 18 IU/ml (Omicron) for mRNA-1273 recipients), compared to the AZD1222 recipients (GMT IC_50_ of 17 (Alpha), 10 (Beta), 13 (Gamma), 10 (Delta), <2 IU/ml (Omicron)), and the Ad26.COV2.S recipients (GMT IC_50_ of 11 (Alpha), 10 (Beta), 11 (Gamma), 14 (Delta), <2 IU/ml (Omicron)). The fold reduction in VOCs neutralization compared to WT was similar for all groups and consistent with previous reports for convalescent sera and vaccine sera showing the largest decrease of neutralization capacity against the Omicron variant (9- to 80-fold; Figure S3B), followed in order by Beta, Delta, Gamma and Alpha. However, the decrease of neutralization for the Omicron variant was lower for the mRNA-1273 recipients compared to the BNT162b2 recipients, which resulted in significantly higher Omicron neutralization titers for mRNA-1273 recipients compared to the other groups (Figure 2B). Overall, binding and neutralizing antibody responses correlated very well for WT and VOCs (r= 0.7995, p<0.001 for wild-type; Figure S3A-B). Antibody binding titers against the VOCs S proteins on the other hand were largely unaffected (Figure S2A), suggesting that neutralizing antibodies form a minority among all antibodies.

**Figure 2:**
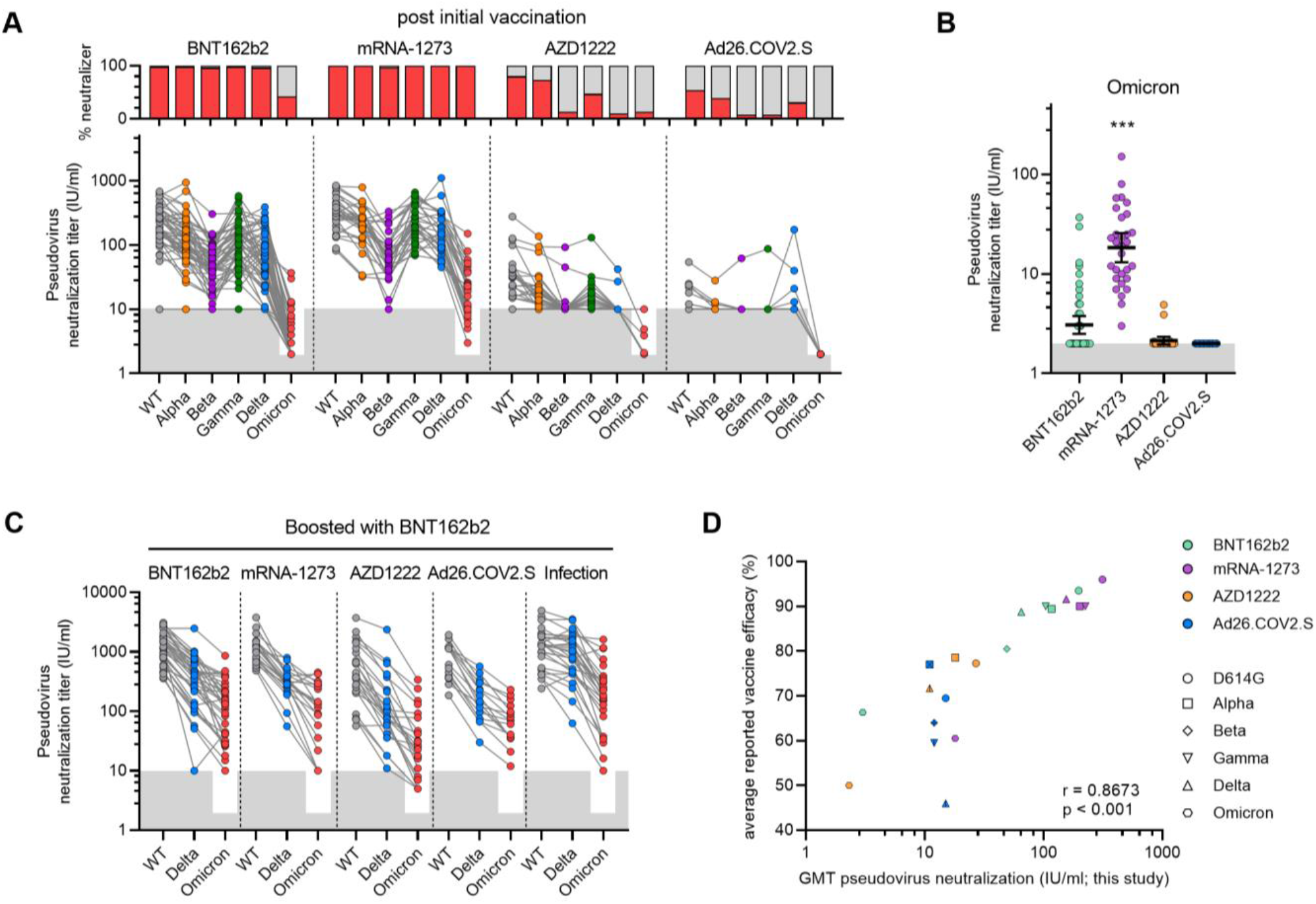
Binding and neutralization titers post-vaccination against VOCs. (A) Paired neutralization IC_50_ titers (UI/ml) of D614G and VOCs pseudoviruses for sera collected post initial vaccination series for the four vaccination groups (lower panel). The lower cut-off for neutralization was set at an IC_50_ of 10 or for Omicron at 2 IU/ml (grey shading). Percentage vaccinee with detectable neutralization titers in red (upper panel). (B) Neutralization titers of Omicron pseudovirus for sera post initial vaccination series. Geometric mean titer (GMT) and 95% CI are indicated. The lower cutoff for neutralization was set at an IC_50_ of 2 IU/ml (grey shading). *** p < 0.001 non-parametric Kruskal-Wallis test for mRNA-1273 compared to the other groups. (C) Paired neutralization IC_50_ titers (IU/ml) of D614G and VOCs pseudoviruses for sera collected post booster vaccination for the four vaccination groups and convalescent group (RECoVERED cohort; four weeks after single dose BNT162b2 vaccination up to 15 months post infection, n=28). The lower cut-off for neutralization was set at an IC_50_ of 10 or for Omicron at 2 IU/ml (grey shading). Uni- and multivariable linear regression analysis results in Table S4. (D) GMT IC_50_ neutralization titers of D614G and VOCs plotted against the average of reported vaccine efficacy against symptomatic infection (Table S2). Vaccine groups are indicated by colors with BNT162b2 in green, mRNA-1273 in purple, AZD1222 in orange and Ad26.COV2.S in blue. Circles represent WT data, squares for Alpha, diamond for Beta, nabla triangle for Gamma and delta triangle for Delta. Crossed symbols are at the neutralization cutoff. Spearman’s rank correlation coefficient with p-value are indicated.

Importantly, the proportion of individuals who did not show detectable VOC neutralization after completing the initial vaccination series was substantial in the AZD1222 and Ad26.COV2.S recipients (AZD1222 recipients: 8/30 non-responders for Alpha, 26/30 for Beta, 16/30 for Gamma, 27/30 for Delta, and 30/30 when considering <10 IU/ml and 26/30 when considering <2 IU/ml for Omicron; Ad26.COV2.S recipients 8/13 non-responders (Alpha), 12/13 (Beta), 12/13 (Gamma), 9/13 (Delta), and 13/13 (<2 IU/ml; Omicron) versus BNT162b2 recipients: 1/50 non-responders (Alpha), 2/50 (Beta), 1/50 (Gamma), 2/50 (Delta), and 46/50 (<10 IU/ml) or 29/50 (<2 IU/ml; Omicron) and mRNA-1273 recipients: 1/30 non-responders (Alpha), 2/30 (Beta), 0/30 (Gamma), 0/30 (Delta), and 9/30 (<10 IU/ml) or 0/30 (<2 IU/ml; Omicron); Figure 2A).

Four weeks after BNT162b2 booster vaccination the neutralization titers against VOCs (Delta and Omicron) were significantly increased, with all participants showing detectable neutralization titers >2 IU/ml and only 6/13 AZD1222 recipients between 2 – 10 IU/ml against Omicron (Figure 2C). Overall, after the initial vaccination series, the mRNA vaccines induced substantial levels of neutralizing antibodies against currently defined VOCs, with the exception of Omicron, while the adenovirus vector-based vaccines were much less efficient in doing so against all VOCs. However, the BNT162b booster vaccination subsequently induced detectable VOC neutralization responses in all recipients. The GMT binding and neutralization titers from our study correlated strongly with the levels of protection from symptomatic infection by the respective strains as obtained from vaccine efficacy studies (r=0.9235, p<0.001; r=0.8673, p<0.001, respectively; Figure 2D, S3C and Table S2 [2,3,5,14–18,20,22–24,36–52]), reinforcing the association between neutralization and protection from infection [19,55,56].

### Neutralizing antibody responses against VOI

Finally, we evaluated neutralization of a number of variants of interest (VOI) and other SARS-CoV-2 variants, including Kappa (B.1.617.1), Lambda (C.37), Epsilon (B.1.429), Iota (B.1.526), Zeta (B.1.1.28.P.2/P.2), Mu (B.1.621) [57], as well as subvariants within the Alpha and Beta VOC lineages (Table S1), after completion of the initial vaccination series. As sera quantities were limited, we tested these viruses against pooled sera from each vaccine group, as well as pooled sera from two convalescent cohorts, the COSCA and RECOVERED [32,58]. Since most Ad26.COV2.S recipients had undetectable neutralizing ability against VOC’s, these sera were not included in this analysis. Furthermore, a selection of monoclonal antibodies (Mab) isolated from participants of the COSCA cohort were also tested against the different VOCs and VOIs (COVA1-16, COVA1-18 and COVA2-15 against the RBD; COVA1-22 and COVA2-17 against the NTD; and COVA1-25 against an unknown epitope) [59].

We included the five VOCs in this analysis and found that the neutralization IC_50_ values obtained with the pooled sera were highly concordant with the GMT IC_50_ values of the individual sera, indicating that the pooling of sera yields representable results. The set of pooled sera had diverse neutralization titers against the VOCs and VOIs (Figure 3A). In particular, the Beta, Omicron, Kappa and Mu variants showed reduced sensitivity to neutralization (Figure S3D), confirming previously observed fold reductions [19,60]. The rank order between the different vaccines was consistent between the various VOCs and VOIs. The additional E484K mutation in the Alpha variant caused an additional 2.8-fold reduction in neutralization for all pools, corroborating the impact of this single RBD mutation on neutralization. The Beta, Gamma, Iota, Zeta and Mu variants have this mutation, while the Kappa variant has the E484Q and Omicron the E484A mutation, contributing to their reduced sensitivity. The differences observed between these VOCs and VOIs indicate that other mutations in addition to E484K/Q/A, such as the K417T/N and L452R/Q in the RBD and mutations in the N-Terminal Domain (NTD), contribute to decreased sensitivity to neutralization.

**Figure 3:**
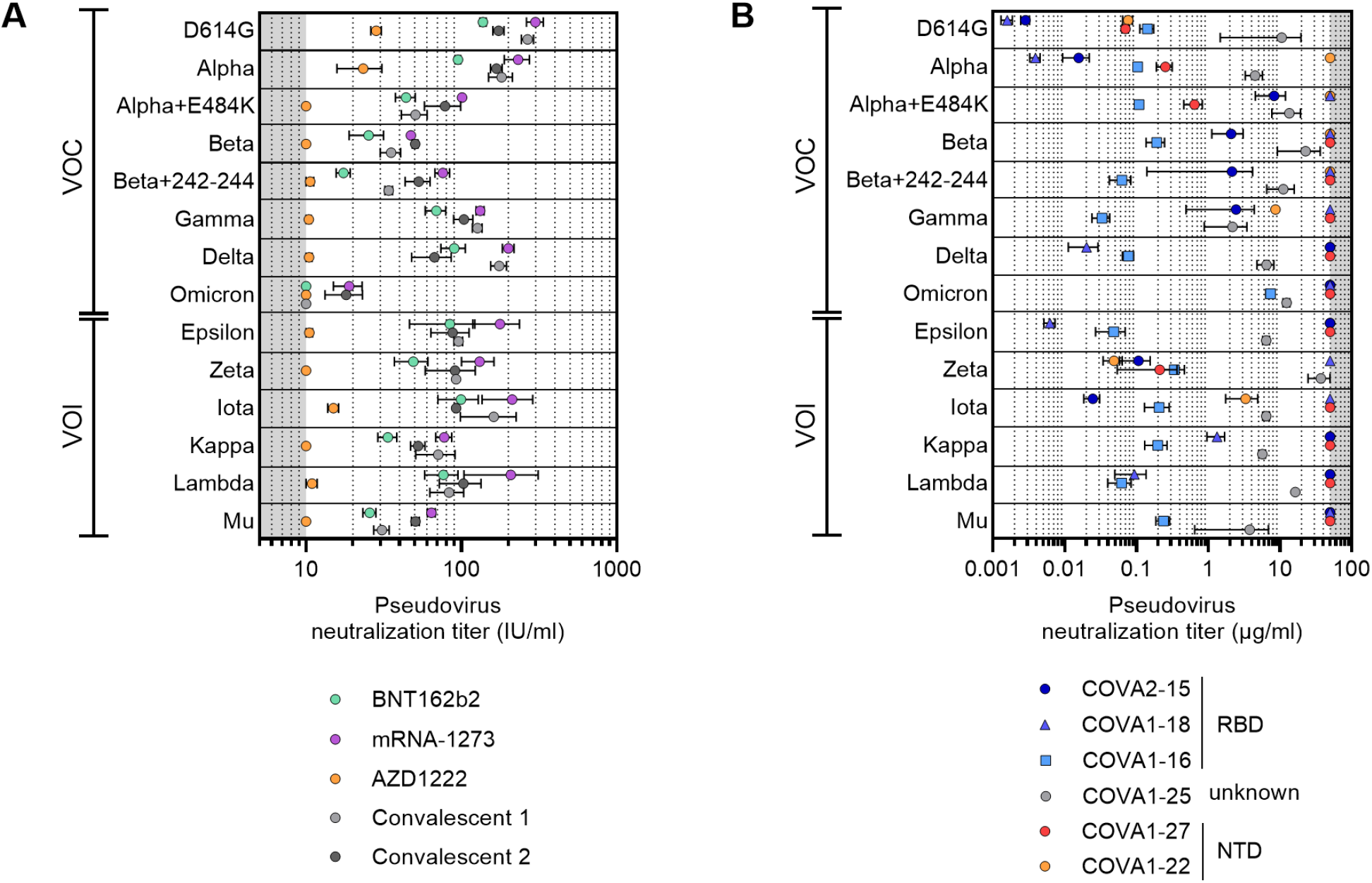
Neutralization titers of pooled sera and monoclonal antibodies against VOCs and VOIs. (A) Neutralization IC_50_ titers (IU/ml) of SARS-CoV-2 variant pseudoviruses for pooled sera for the vaccination groups (excluding the Ad26.COV2.S group) after completing initial vaccination series. The lower cutoff for neutralization was set at an IC_50_ of 10 IU/ml (grey shading). Convalescent group 1 (light gray) consists of pooled COSCA sera representing COVID-19 patients between 4-6 weeks post symptom onset and convalescent group 2 (dark gray) consists of pooled RECoVERED sera representing COVID-19 patients up to seven months post symptom onset (median three months), who experienced mild to severe COVID-19. Vaccine groups are indicated by colors with BNT162b2 in green, mRNA-1273 in purple and AZD1222 in orange. All data points shown here represent mean and standard deviation of at least three replications. (B) Half-maximal inhibitory concentrations (IC_50_) of SARS-CoV-2 variant pseudoviruses neutralization for monoclonal antibodies isolated from COSCA participants [59]. The cutoff for neutralization was set at an IC_50_ of 50 μg/ml (grey shading). All data points shown here represent mean and standard deviation of at least three replications.

To further elucidate the effect of these mutations on antibody potency, we tested several potent neutralizing antibodies targeting the RBD or NTD. The RBD targeting MAb COVA1-16 was able to neutralize all VOCs and VOIs with IC_50_ of 0.020 – 0.2 μg/ml, except Omicron at 6.4 μg/ml, consistent with its previously established insensitivity to mutations in emerging SARS-CoV-2 variants and the Omicron mutations S371L and S375F in its epitope [32,61–64] (Figure 3B). The weakly neutralizing non-RBD MAb COVA1-25, with an unknown target epitope, also neutralized all VOC and VOI tested, albeit with some variation in potency. In contrast, RBD-targeting COVA1-18 was strongly active against the Alpha, Delta, Epsilon and Lambda variants and also neutralized Kappa, but was unable to neutralize the Beta, Gamma, Omicron, Iota, Zeta and Mu variants. These data suggest that COVA1-18 neutralization is knocked out by the E484K/A mutation and this was confirmed by the comparison of Alpha (COVA1-18 sensitive) and Alpha + E484K (COVA1-18 insensitive). COVA1-18 activity against the Kappa variant suggests that the antibody is able to accommodate the E484Q mutation. Another RBD MAb, COVA2-15 showed reduced activity against most VOCs and VOIs and lost neutralization against Delta, Omicron, Kappa, Lambda, Epsilon and Mu, which all, except Omicron and Mu carry a mutation at residue 452, either L452R or L452Q, suggesting that L452 is important for COVA2-15 neutralization, however also other mutations affect COVA2-15 activity. The NTD MAbs COVA1-22 and COVA2-17 showed reduced or loss of activity against all variants except Zeta that does not have any mutations in the NTD. As the NTD mutations are very diverse between the different variants, it is not possible to predict the most important residues for neutralization for COVA1-17 and COVA1-22.

## Discussion

Current and future SARS-CoV-2 variants could potentially jeopardize the effectiveness of vaccines in curbing the pandemic by escaping vaccine-induced immune responses. We present a direct comparison of the ability of four approved SARS-CoV-2 vaccines to induce neutralizing antibodies against VOCs, revealing that the mRNA vaccines are profoundly superior to the adenovirus vector-based vaccines at inducing neutralizing antibodies. We further show that the antibodies in SARS-CoV-2 vaccine recipients, sampled around the expected peak of their immunity, showed a marked decrease in neutralization potency against the VOCs, especially the Omicron variant, which was shown to form a separate antigenic cluster [65]. When neutralization activity against the original strain was limited, as observed after AZD1222 or Ad26.COV2.S vaccination, the capability to potently neutralize different variants is severely diminished. An mRNA booster significantly improved the neutralizing ability, including against the currently circulating Omicron variant.

The differences between mRNA and adenovirus vector-based vaccines might have several reasons. First, Ad26.COV.2 was only used as a single dose whilst a second boost immunization might very well enhance its ability to induce neutralizing antibodies. A recent study suggests that this might indeed be the case [66]. This argument does not hold for AZD1222 as the increase of the antibody levels after the second dose was substantially less pronounced in the AZD1222 recipients compared to the mRNA vaccine recipients. The AZD1222 vaccine encodes for an unmodified S protein, while Ad26.COV.S, mRNA-1273 and BNT162b2 vaccines s encode for a proline-stabilized version of S, which might be more conducive for the induction of neutralizing antibodies [67]. Other platform-intrinsic factors might also play a role, such as differences in S expression levels and/or the duration of S expression. Interestingly, the Ad26.COV2.S recipients showed no decline of neutralization titers over a 6 month period, while the BNT162b2, mRNA-1273 and AZD1222 vaccines showed similar substantial decline in antibody titers. Further studies are warranted to investigate the underlying reasons.

Our neutralization results correlated remarkably well with reported vaccine efficacy of the four vaccines against VOCs (r=0.8673, p<0.001) and reinforce the reports that neutralizing antibodies are a strong correlate of protection [19,55,56]. However, strong neutralizing antibody responses do not alone account for the protection by current vaccines [68]. While neutralizing antibody levels were low and often undetectable in our assay after the initial vaccination series with the adenovirus vector-based vaccines in comparison to mRNA vaccines, especially against the VOC, the vaccines still show substantial vaccine efficacy against symptomatic infection and severe disease (>60%), albeit less than the mRNA vaccines [2,3,5,14–18,20,22–24,36–52]. This strongly suggests that other immune components play important roles. These include low levels of neutralizing antibodies (IC_50_ <10), T cells, and possibly non-neutralizing antibodies with effector functions [69–72]. Furthermore, memory B cell responses are likely to play a role, in particular in protection against severe disease [73,74]. An additional vaccine administration to AZD1222 and Ad26.COV.2 recipients, either with the same vaccine or with an mRNA vaccine, could further boost this protection. Recent studies indeed suggest that booster vaccines and heterologous adenovirus prime mRNA boost regimens might be superior to adenovirus only or mRNA only [75,76]. Indeed, we observed significant increases in antibody titers after the heterologous BNT162b2 boost vaccination.

After one vaccine dose, we observed higher neutralization titers for mRNA-1273 recipients compared to the individuals receiving BNT162b2. Another study also reported that mRNA-1273 was slightly more efficient at inducing neutralizing antibodies compared to BNT162b2 [77]. Furthermore, mRNA-1273 induced significantly higher neutralizing antibody titers against Omicron than BNT162b2. One explanation could be the higher mRNA dose in the mRNA-1273 vaccine (100 μg versus 30 μg in the BNT162b2 vaccine). This might also explain the reported limited efficacy of the Curevac vaccine (CVnCoV), which contained only 12 μg of mRNA, although the instability of the mRNA due to the use of unmodified bases might have contributed to this as well [78].

Six to nine months after the initial vaccination series, binding and neutralization titers were still considerably lower in the adenovirus-vector-based vaccines, despite the relative durability of titers in the Ad26.COV.2 recipients. However, the observation that the inferior response can be salvaged by administration of an mRNA booster is encouraging. However, this salvation is only partial: post-booster titers remain lowest in those initially vaccinated with adenovector-based vaccines. This may leave these individuals less well protected, as suggested by the strong correlation between neutralization titers and reported vaccine efficacy, although it is yet uncertain to what extent this is the case, and whether this can be solved by subsequent boosters.

Our results, as well as many previous studies, identify the main culprits among the mutations present in VOCs and VOIs for reducing neutralization sensitivity. As RBD antibodies dominate the neutralizing antibody response, RBD mutations proved critical. E484K/A (present in Beta, Gamma and Omicron) abrogates sensitivity to a number of RBD antibodies, while L452R (present in Delta) and K417N/T (present in Beta, Gamma and Omicron) affect other subsets of RBD antibodies [60,61,79]. Several therapeutic antibodies currently in use for COVID-19 treatment are affected by these mutations and have reduced activity against VOCs [80]. The accumulation of these mutations, as well as others, in Omicron and in the context of a heavily mutated lab-built version of the Delta variant, renders them profoundly more resistant to neutralization [81].

There are several limitations of our study. First, our study includes substantially more female than male participants, reflecting the gender distribution among HCW at our institute. Second, the age distribution in the four groups is not identical. In particular, the AZD1222 group is considerably older as a consequence of restrictive use of the AZD1222 vaccine in individuals aged 60-64 years in the Netherlands. As immune responses tend to become weaker with higher age, this is a relevant factor when considering the weaker responses in the AZD1222 group. Reassuringly, our findings were robust to adjustment for participant sex and age. Finally, we did not include long-term measurements post-booster. It will be relevant to study the durability of the neutralizing antibody responses after mRNA booster vaccination after initial vaccination with each of these vaccines.

We have analyzed known VOCs and many VOIs. While we cannot predict how our results apply to future variants, the data with Omicron reveal how antigenic drift can substantially impact the extent to which vaccine-induced responses can cross-neutralize new antigenic variants. Current VOCs up to Omicron were probably selected based on increased fitness and/or transmissibility, while Omicron and future variants may very well be selected based on escape from immunity when more and more people are vaccinated or have experienced COVD-19. Such escape variants are more resistant to neutralizing antibodies induced by current vaccines, especially when neutralization titers are low for the adenovirus vector-based vaccines and should prompt boosting and vaccine updates based on circulating variants.

The implication of our results is that individuals receiving one of the adenovirus vector-based vaccines are more vulnerable to infection with the VOCs, which is consistent with the lower efficacy of these vaccines against symptomatic infection with VOCs compared to the mRNA vaccines, although all vaccines are highly effective at preventing severe disease by VOCs. While circulating antibodies might be unable to neutralize such emerging viruses, memory B cells are still likely to recognize them and undergo new rounds of affinity maturation, resulting in new neutralizing antibodies that should kick-in in time to prevent severe disease after infection.

## Data Availability

All data are available in the main text or supplementary materials. Reagents used in this study are available upon reasonable request under an MTA with Amsterdam UMC.

## Acknowledgements

We thank Dr. Paul Bieniasz and Theodora Hatziioannou of the Howard Hughes Medical Institute, Rockefeller University, New York, USA and Dr. Beatrice Hahn of the Perelman School of Medicine, University of Pennsylvania, Philadelphia, USA for donating cells and reagents for pseudovirus neutralization assays; Dirk Eggink and Chantal Reusken of the National Institute for Public Health and the Environment, Bilthoven, the Netherlands for providing the SARS-CoV-2 Delta and Omicron S proteins; Johan Reimerink, Fion Brouwer, Marieke Hoogerwerf and Tarek Munawar, Bas J. Verkaik, Orlane J.A. Figaroa, Peter J. de Vries, Tessel M. Boertien, Neeltje A. Kootstra and all researchers, nurses and students of the RECoVERED Study team for technical assistance; and all the participants of the S3/HCW, COSCA and RECoVERED studies.

## Author contributions

Conceptualization: M.J.v.G., J.J.S., M.K.B., R.W.S.; Funding acquisition: M.J.v.G., M.D.d.J., J.J.S., M.K.B., R.W.S.; Investigation: M.J.v.G., A.H.A.L., K.v.d.S., B.A., I.B., Me.P., J.A.B., M.O., J.H.B., K.T., J.v.R., M.G.; Methodology: M.J.v.G., J.J.S., M.K.B., R.W.S.; Project administration: M.J.v.G., A.H.A.L., K.v.d.S., B.A., J.J.S., M.K.B.; Resources: L.A.V., M.A.S., M.S., E.W., H.D.G.v.W., M.G., J.S., T.G.C., S3/HCW, A.P.J.V., M.D.d.J., G.J.d.B.; Supervision: M.J.v.G., A.P.J.V., Ma.P., M.D.d.J., G.J.d.B., J.J.S., M.K.B., R.W.S.; Writing – original draft: M.J.v.G., R.W.S.; Writing – review & editing: all authors.

## Notes

### Competing Interest Statement

Amsterdam UMC filed a patent application on SARS-CoV-2 monoclonal antibodies including the ones used in this manuscript.

### Funding Statement

the Netherlands Organization for Scientific Research (NWO) ZonMw (no. 10430022010023 no. 10150062010002 no. 91818627)

the Bill & Melinda Gates Foundation (no. INV-002022 no. INV008818 no. INV-024617)

the Amsterdam UMC through the AMC Fellowship and the Corona Research Fund

the European Unions Horizon 2020 program (no. 101003589).

### Author Declarations

The S3 study, the COSCA study and the RECoVERED study were approved by the medical ethical review board of the Amsterdam University Medical Centers (NL73478.029.20, NL73281.018.20 and NL73759.018.20, respectively). All participants provided written informed consent.

### Summary of Updates

This version of the manuscript has been revised to include booster vaccination and monoclonal antibodies data.

