## Supplemental Materials for "Antibody responses against SARS-CoV-2 variants induced by four different SARS-CoV-2 vaccines"

**This PDF file includes:**

Figure S1 to S3

Table S1 to S5

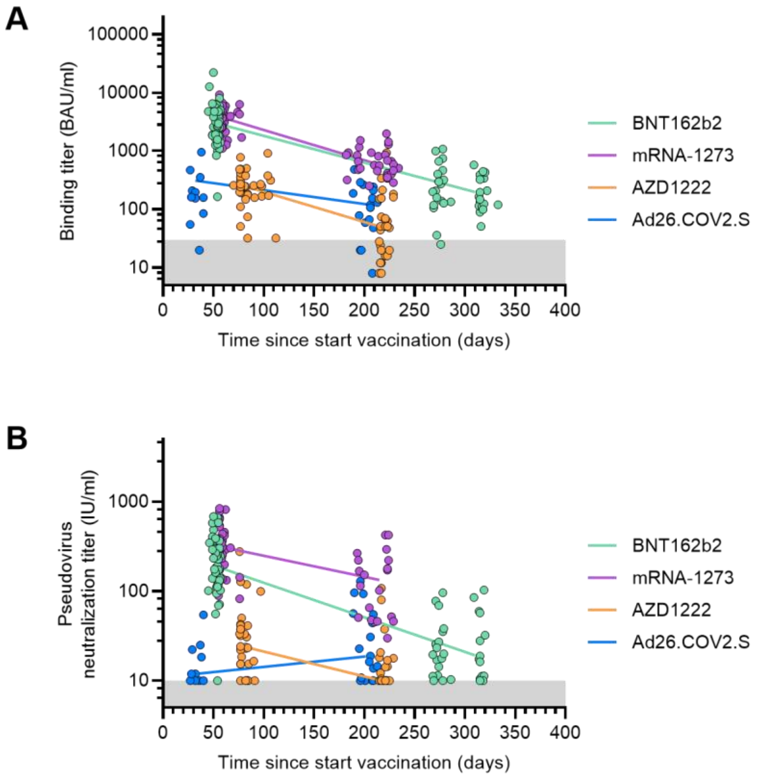


**Figure S1: Binding and neutralization titer decline.** Binding titers to wild-type S protein (BAU/ml) of 1:100,000 diluted sera (A) and neutralization titers (IU/ml) of D614G pseudovirus (B) collected post complete vaccination and pre-booster for the four vaccination groups. The lower cutoff for binding was set at 30 BAU/ml (grey shading) for binding titers and at 2 IU/ml for neutralization titers. Lines connect the GMT of four weeks post complete vaccination and GMT of pre-booster titers per group. Vaccine groups are indicated by colors with BNT162b2 in green, mRNA-1273 in purple and AZD1222 in orange.

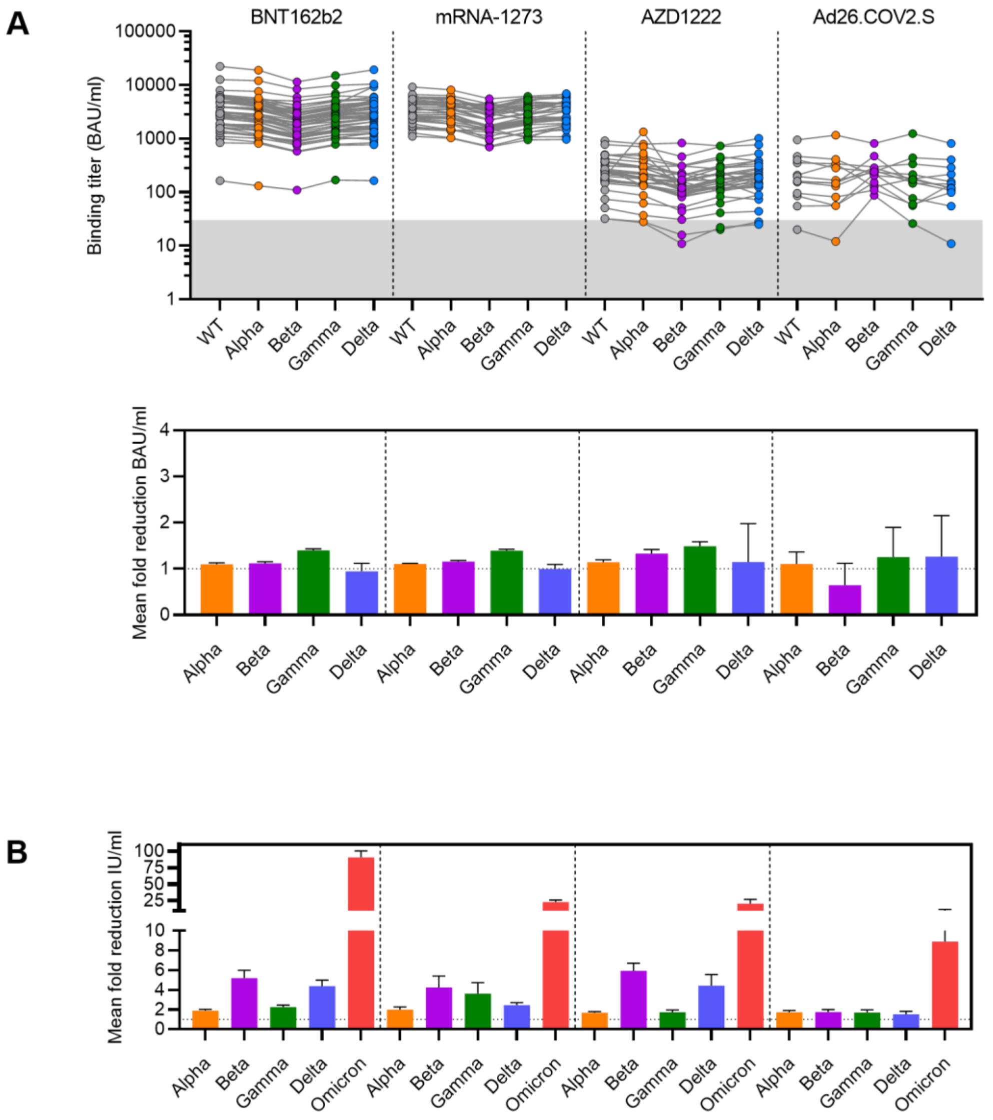


**Figure S2: Binding and neutralization titers post-vaccination against the VOCs.** (A) Paired binding titers to wild-type and VOCs S protein (BAU/ml) of 1:100,000 diluted sera collected post complete vaccination for the four vaccination groups (upper plot). The lower cut-off for binding was set at a 30 BAU/ml (grey shading). Mean ± SEM fold reductions in binding titers against the VOCs in comparison to wild-type (lower plot). (B) Mean ± SEM fold reductions in neutralization $IC_{50}$ titers against the VOCs pseudoviruses in comparison to neutralization $IC_{50}$ titers against D614G pseudovirus.

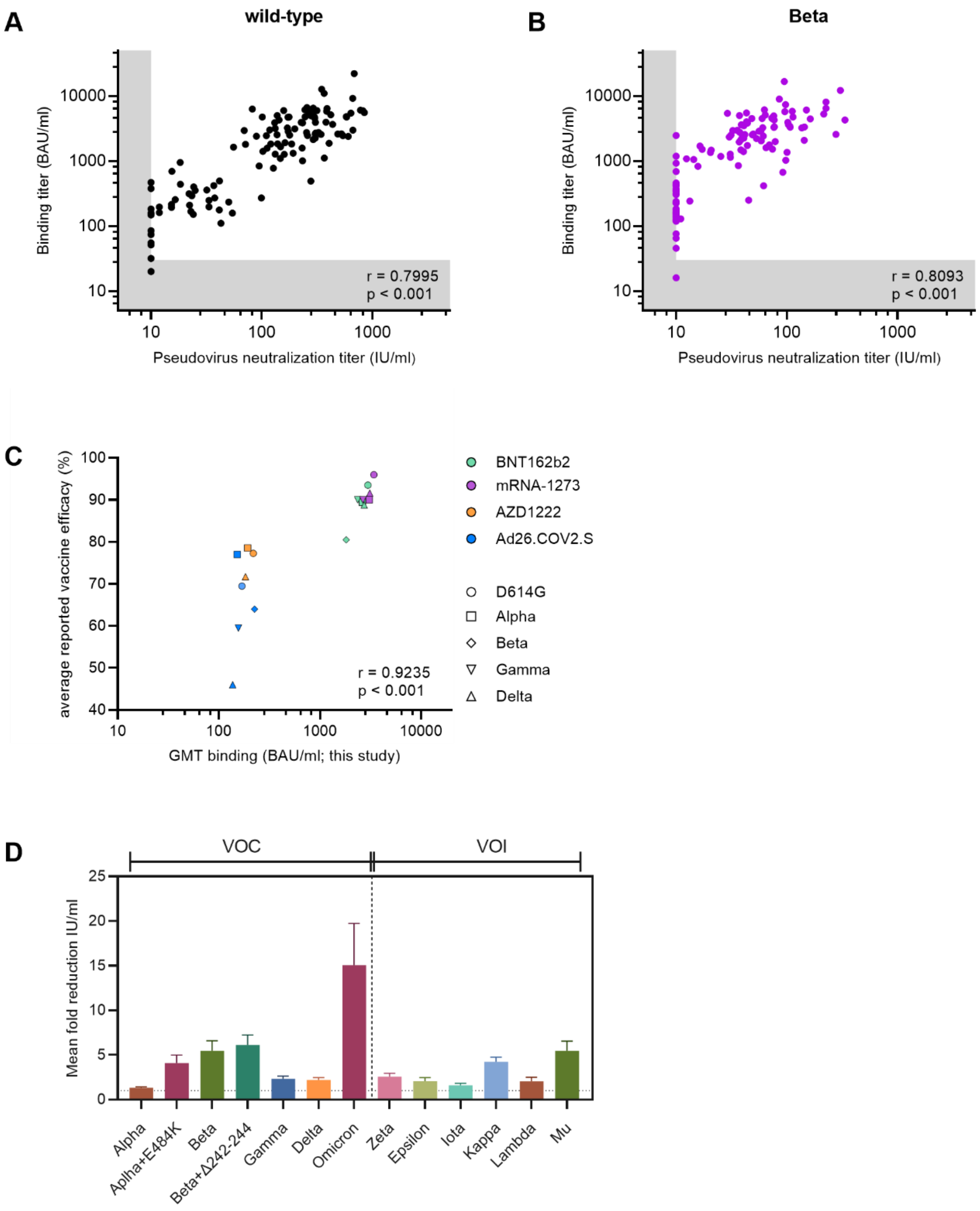


**Figure S3: Binding and neutralization post-vaccination titers against VOCs correlated.** Correlation between wild-type (A) and Beta (B) binding (BAU/ml) and neutralization titers (IU/ml). Spearman's rank correlation coefficients with p-values are indicated. (C) GMT (BAU/ml) of wild-type and VOCs plotted against the average reported vaccine efficacy against symptomatic infection (Table S4). Vaccine groups are indicated by colors with BNT162b2 in green, mRNA-1273 in purple, AZD1222 in orange and Ad26.COV2.S in blue. Circles represent WT data, squares for Alpha, diamond for Beta, nabla triangle for Gamma and delta triangle for Delta. Spearman's rank correlation coefficients with p-values are indicated. (D) Mean ± SEM fold reductions in neutralization $IC_{50}$ titers for the serum pools combined against the VOCs and VOIs pseudoviruses in comparison to $IC_{50}$ titers against D614G pseudovirus.

**Table S1: Protein and pseudovirus S constructs contain the following mutations compared to the WT (Wuhan Hu-1; GenBank: MN908947.3).**

| VOC S proteins | | | | | | |
|---|---|---|---|---|---|---|
| **Alpha**<br>B.1.1.7 | **Beta**<br>B.1.351 | **Gamma**<br>P.1 | **Beta**<br>B.1.351 | **Omicron**<br>B.1.1.529 | | |
| Δ69-70 | L18F | L18F | T19R | A67V | S375F | Y505H |
| Δ144 | D80A | T20N | G142D | Δ69-70 | K417N | T547K |
| N501Y | D215G | P26S | E156G | T95I | N440K | D614G |
| A570D | Δ242-244 | D138Y | Δ157-158 | G142D | G446S | H655Y |
| D614G | K417N | R190S | L452R | Δ143-145 | S477N | N679K |
| P681H | E484K | K417T | T478K | Δ211 | T478K | P681H |
| T716I | N501Y | E484K | D614G | L212I | E484A | N764K |
| S982A | D614G | N501Y | P681R | ins214EPE | Q493K | D796Y |
| D1118H | A701V | D614G | D950N | G339D | G496S | N856K |
| | | H655Y | | S371L | Q498R | Q954H |
| | | T1027I | | S373P | N501Y | |

| | VOC pseudovirus | | | | | | | |
|---|---|---|---|---|---|---|---|---|
| **D614G**<br>B.1 | **Alpha**<br>B.1.1.7 | **Alpha +E484K**<br>B.1.1.7 | **Beta**<br>B.1.351 | **Beta +242-244**<br>B.1.351 | **Gamma**<br>P.1 | **Delta**<br>B.1.617.2 | **Omicron**<br>B.1.1.529 BA.1 | |
| D614G | Δ69-70 | Δ69-70 | L18F | L18F | L18F | T19R | A67V | Q498R |
| | Δ144 | Δ144 | D80A | D80A | T20N | G142D | Δ69-70 | N501Y |
| | N501Y | E484K | D215G | D215G | P26S | E156G | T95I | Y505H |
| | A570D | N501Y | Δ242-244 | L242H | D138Y | Δ157-158 | G142D | T547K |
| | D614G | A570D | K417N | R246I | R190S | L452R | Δ143-145 | D614G |
| | P681H | D614G | E484K | K417N | K417T | T478K | Δ211 | H655Y |
| | T716I | P681H | N501Y | E484K | E484K | D614G | L212I | N679K |
| | S982A | T716I | D614G | N501Y | N501Y | P681R | ins214EPE | P681H |
| | D1118H | S982A | A701V | D614G | D614G | D950N | G339D | N764K |
| | | D1118H | | A701V | H655Y | | S371L | D796Y |
| | | | | | T1027I | | S373P | N856K |
| | | | | | | | S375F | Q954H |
| | | | | | | | K417N | N969K |
| | | | | | | | N440K | L981F |
| | | | | | | | G446S | |
| | | | | | | | S477N | |
| | | | | | | | T478K | |
| | | | | | | | E484A | |
| | | | | | | | Q493K | |
| | | | | | | | G496S | |

| VOI pseudovirus | | | | | |
|---|---|---|---|---|---|
| **Epsilon**<br>B.1.429 | **Zeta**<br>P.2 | **Iota**<br>B.1.526 | **Kappa**<br>B.1.617.1 | **Lambda**<br>C.37 | **Mu**<br>B.1.621 |
| S13I | E484K | L5F | T95I | G75V | Δ69-70 |
| W152C | D614G | T95I | G142D | T76I | T95I |
| L452R | V1176F | D253G | E154K | Δ246-252 | Y144S |
| D614G | | E484K | L452R | D253N | Y145N |
| | | D614G | E484Q | L452Q | R346K |
| | | A701V | D614G | F490S | E484K |
| | | | P681R | D614G | N501Y |
| | | | Q1071H | T859N | A570D |
| | | | | | D614G |
| | | | | | P681H |
| | | | | | T716I |
| | | | | | D950N |
| | | | | | S982A |

**Table S2: Reported vaccine efficacy (References; [2,3,5,14–18,20,22-24,36–52]).**

| Vaccine | VOC | VE | Reference |
|---|---|---|---|
| BNT162b | WT | 96% | Thomas et al |
| BNT162b | WT | 95% | Polack et al |
| BNT162b | WT | 91% | Chung et al |
| BNT162b | WT | 92% | Nasreen et al |
| BNT162b | Alpha | 68% | Meyer et al |
| BNT162b | Alpha | 89% | Pilishvili et al |
| BNT162b | Alpha | 98% | Glatman-Freedman et al |
| BNT162b | Alpha | 95% | Andrews et al 2022-2 |
| BNT162b | Alpha | 97% | Katz et al |
| BNT162b | Alpha | 87% | Kissling et al |
| BNT162b | Alpha | 94% | Lopez Bernal et al |
| BNT162b | Alpha | 82% | Martínez-Baz et al |
| BNT162b | Alpha | 90% | Public Health England |
| BNT162b | Alpha | 97% | Angel et al |
| BNT162b | Alpha | 90% | Abbu-Raddad et al |
| BNT162b | Alpha | 88% | Nasreen et al |
| BNT162b | Alpha | 92% | Sheikh et al |
| BNT162b | Alpha | 73% | Sheikh et al |
| BNT162b | Beta | 75% | Abbu-Raddad et al |
| BNT162b | Beta | 86% | Nasreen et al |
| BNT162b | Gamma | 90% | Nasreen et al |
| BNT162b | Delta | 90% | Prunas et al |
| BNT162b | Delta | 87% | Kissling et al https://osf.io/3nhps/ |
| BNT162b | Delta | 93% | Powell et al |
| BNT162b | Delta | 91% | Andrews et al 2022-1 |
| BNT162b | Delta | 93% | Reis et al |
| BNT162b | Delta | 92% | Nordstrom et al |
| BNT162b | Delta | 83% | Andrews et al 2022 |
| BNT162b | Delta | 44% | Tang et al |
| BNT162b | Delta | 88% | Lopez Bernal et al |
| BNT162b | Delta | 92% | Nasreen et al |
| BNT162b | Omicron | 62% | Chemaitelly et al |
| BNT162b | Omicron | 71% | Powell et al |
| BNT162b | Omicron | 66% | Andrews et al 2022-2 |
| mRNA-1273 | WT | 94% | Baden et al |
| mRNA-1273 | WT | 94% | Chung et al |
| mRNA-1273 | WT | 98% | Nasreen et al |
| mRNA-1273 | Alpha | 96% | Pilishvili et al |
| mRNA-1273 | Alpha | 92% | Nasreen et al |
| mRNA-1273 | Delta | 98% | Kissling et al https://osf.io/3nhps/ |
| mRNA-1273 | Delta | 95% | Andrews et al 2022-1 |
| mRNA-1273 | Delta | 96% | Nordstrom et al |
| mRNA-1273 | Delta | 95% | Andrews et al 2022-2 |
| mRNA-1273 | Delta | 74% | Tang et al |
| mRNA-1273 | Delta | 94% | Nasreen et al |
| mRNA-1273 | Omicron | 45% | Chemaitelly et al |
| mRNA-1273 | Omicron | 76% | Andrews et al 2022-2 |
| AZD1222 | WT | 62% | Voysey et al |
| AZD1222 | Alpha | 82% | Andrews et al 2022-2 |
| AZD1222 | Alpha | 78% | Hitchings et al |
| AZD1222 | Alpha | 75% | Lopez Bernal et al |
| AZD1222 | Alpha | 89% | Public Health England |
| AZD1222 | Alpha | 87% | Nasreen et al |

| | | | |
|---|---|---|---|
| AZD1222 | Delta | 72% | Kissling et al https://osf.io/3nhps/ |
| AZD1222 | Delta | 83% | Andrews et al 2022-1 |
| AZD1222 | Delta | 68% | Nordstrom et al |
| AZD1222 | Delta | 64% | Andrews et al 2022-2 |
| AZD1222 | Delta | 67% | Lopez Bernal et al |
| AZD1222 | Delta | 88% | Nasreen et al |
| AZD1222 | Delta | 79% | Sheikh et al |
| AZD1222 | Delta | 60% | Sheikh et al |
| AZD1222 | Omicron | 50% | Andrews et al 2022-2 |
| Ad26.COV2.S | WT | 67% | Sadoff et al |
| Ad26.COV2.S | Alpha | 77% | Martínez-Baz et al |
| Ad26.COV2.S | Beta | 64% | Sadoff et al |
| Ad26.COV2.S | Gamma | 51% | Ranzani et al |
| Ad26.COV2.S | Delta | 50% | Kissling et al https://osf.io/3nhps/ |
| Ad26.COV2.S | Delta | 42% | Martínez-Baz et al |

**Table S3: Number of participants included in the binding or neutralization assay per time point per vaccine group.**

| | | Binding | Neutralization |
|---|---|---|---|
| BNT162b2 | pre-vac | 50 | 0 |
| | post-V1 | 46 | 45 |
| | post-V2 | 50 | 50 |
| | +9m V2 | 42 | 36 |
| | post-V3 | 37 | 34 |
| mRNA-1273 | pre-vac | 25 | 0 |
| | post-V1 | 40 | 30 |
| | post-V2 | 39 | 29 |
| | +6m V2 | 31 | 24 |
| | post-V3 | 28 | 20 |
| AZD1222 | pre-vac | 19 | 0 |
| | post-V1 | 42 | 34 |
| | post-V2 | 34 | 30 |
| | +6m V2 | 32 | 29 |
| | post-V3 | 26 | 23 |
| Ad26.COV2.S | pre-vac | 7 | 0 |
| | post-V1 | 13 | 13 |
| | +2m V1 | 13 | 13 |
| | +7m V1 | 19 | 17 |
| | post-V2* | 18 | 16 |

### Table S4: Uni- and multivariable linear regression analysis.

**Geometric means of binding levels against S wild type, BAU/ml (95% CI)**

| | **Post 1 vaccination** | | **Post complete initial vaccination±** | | **Pre-booster** | | **Post-booster** | |
|---|---|---|---|---|---|---|---|---|
| | **Univariable** | **Multivariable¥** | **Univariable** | **Multivariable¥** | **Univariable** | **Multivariable¶** | **Univariable** | **Multivariable¥** |
| BNT162b2 | 417 (324-536) | 394 (280-554) | 2967 (2419-3638) | 2603 (1958-3460) | 192 (138-268) | 340 (120-964) | 2840 (2321-3475) | 2756 (2071-3668) |
| mRNA-1273 | 875 (666-1150)▫▫▫ | 781 (512-1191)▫▫▫ | 3461 (2739-4374) | 2857 (1973-4137) | 595 (404-876)▫▫▫ | 386 (211-705) | 3138 (2488-3958) | 2993 (2097-4271) |
| AZD1222 | 79 (61-103)*** | 95 (60-150)*** | 219 (171-281)*** | 202 (136-301)*** | 37 (25-54)*** | 27 (13-55)*** | 1487 (1117-1892)*** | 1480 (997-2196)▫▫ |
| Ad26.COV2.S | 169 (105-271)▪▪ | 169 (96-296)◦◦ | 169 (113-252)*** | 146 (91-234)*** | 107 (65-175)** | 90 (49-165)* | 1541 (1153-2058)*** | 1484 (1029-2140)*** |

* significantly different from AZD1222 and BNT162b2 (p<0.05) and from mRNA-1273 (p<0.001)
** significantly different from AZD1222 (p=0.001) and mRNA-1273 (p<0.001)
*** significantly different from both mRNA vaccines (p<0.001)
▫▫ significantly different from both mRNA vaccines (p<0.01)
ignificantly different from both vector vaccines and BNT162b2 (p<0.001)
▪▪ significantly different from AZD and BNT162b2 (p<0.01) and mRNA-1273 (p<0.001)
◦◦ significantly different from BNT162b2 (p<0.01) and mRNA-1273 (p<0.001)

± complete vaccination is 2 doses of BNT162b2, mRNA-1273 or AZD1222 of 1 dose of Ad26.COV2.S, for Ad26.COV2.S the same time point as post 1 vaccination
¥ corrected for age and sex, geometric means and %95 CI represent median age of the cohort (47 years ) and female gender
¶ corrected for age, sex and time between full vaccination and pre-booster serum sampling, geometric means and %95 CI represent median age of the cohort (47 years ), female gender and median time between full vaccination and pre-booster serum sampling (28 weeks)

**Geometric means of neutralizing antibodies against D614G, $IC_{50}$ in IU/ml (95% CI)**

| | **Post 1 vaccination** | | **Post complete initial vaccination±** | | **Pre-booster** | | **Post-booster** | |
|---|---|---|---|---|---|---|---|---|
| | **Univariable** | **Multivariable¥** | **Univariable** | **Multivariable¥** | **Univariable** | **Multivariable¶** | **Univariable** | **Multivariable¥** |
| BNT162b2 | 16 (12-20) | 15 (10-20) | 197 (160-243) | 152 (113-204) | 24 (18-33) | 42 (15-113) | 1154 (881-1511) | 1192 (817-1738) |
| mRNA-1273 | 32 (24-42)** | 28 (18-43)▫ | 313 (239-409)** | 220 (149-325)▫ | 117 (79-172)▫▫▫ | 92 (53-160) | 1106 (778-1572) | 1160 (717-1878) |
| AZD1222 | 13 (10-18)▪▪▪ | 13 (8-22)▪▪ | 26 (20-34)*** | 19 (12-29)*** | 8 (6-12)*** | 10 (5-19)▪ | 445 (321-617)*** | 379 (221-650)*** |
| Ad26.COV2.S | 14 (9-22)▪▪ | 13 (8-23)▪▪ | 14 (9-21)* | 10 (6-16)* | 21 (13-33)▫▫ | 20 (11-35)▪▪▪ | 700 (473-1037)▫ | 735 (445-1213)◦ |

* significantly different from AZD1222 (p<0.05) and both mRNA vaccines (p<0.001)
** significantly different from BNT162b2 (p<0.01)
*** significantly different from both mRNA vaccines (p<0.001)
▫ significantly different from BNT162b2 (p<0.05)
▫▫ significantly different from AZD1222 (p<0.01) and mRNA-1273 (p<0.001)
▫▫▫ significantly different from BNT162b2 (p<0.001)
▪ significantly different from BNT162b2 (p<0.05) and mRNA-1273 (p<0.001)
▪▪ significantly different from mRNA-1273 (p<0.01)
▪▪▪ significantly different from mRNA-1273 (p<0.001)
◦ significantly different from AZD1222 and BNT162b2 (p<0.05)

± complete vaccination is 2 doses of BNT162b2, mRNA-1273 or AZD1222 of 1 dose of Ad26.COV2.S
¥ corrected for age and sex, geometric means and %95 CI represent median age of the cohort (47 years ) and female gender
¶ corrected for age, sex and time between full vaccination and pre-booster serum sampling, geometric means and %95 CI represent median age of the cohort (47 years ), female gender and median time between full vaccination and pre-booster serum sampling (28 weeks)

**Comparison post initial vaccination series with convalescent cohort (COSCA)**

| Post complete initial vaccination±, univariable | Geometric means of binding levels against S wild type, BAU/ml (95% CI) | Geometric means of neutralizing antibodies against D614G, $IC_{50}$ in IU/ml (95% CI) |
|---|---|---|
| BNT162b2 | 2967 (2212-3979) | 197 (149-260) |
| mRNA-1273 | 3461 (2472-4847) | 313 (218-448)* |
| AZD1222 | 219 (153-313)*** | 26 (18-37)*** |
| Ad26.COV2.S | 169 (95-300)*** | 14 (8-25)*** |
| Convalescent | 3463 (2630-4559) | 175 (138-223)◦◦◦ |

*** significantly different from both mRNA vaccines and convalescent (p<0.001)
* significantly different from BNT162b2 (p=0.047)
◦◦◦ significantly different from both vector vaccines (p<0.001) and mRNA-1273 (0.009)
± complete vaccination is 2 doses of BNT162b2, mRNA-1273 or AZD1222 of 1 dose of Ad26.COV2.S

**Table S5: Individual binding and neutralization titers per vaccine group.**

Can be found in a separate extended data file
